# Engineering a Mobile Platform to Promote Sleep in the Pediatric Primary Care Setting

**DOI:** 10.1101/2020.11.06.20223719

**Authors:** Jonathan A. Mitchell, Knashawn H. Morales, Ariel A. Williamson, Nicholas Huffnagle, Casey Eck, Abigail Jawahar, Lionola Juste, Alexander G. Fiks, Babette S. Zemel, David F. Dinges

## Abstract

**Background:** Pediatricians lack tools to support families at home for the promotion of childhood sleep. We are using the Multiphase Optimization Strategy (MOST) framework to guide the development of a mobile health platform for childhood sleep promotion.

**Purpose:** Under the *preparation phase* of the MOST framework, to demonstrate feasibility of a mobile health platform towards treating children with insufficient sleep.

**Methods:** Children aged 10-12y were enrolled (Study #1: N=30; Study #2: N=43). Participants wore a sleep tracker to measure sleep duration. Data were retrieved by a mobile health platform, programmed to send introductory messages during run-in (2 weeks) and goal achievement messages during intervention (7 weeks) periods. In study #1, participants were randomized to control, gain-framed incentive or loss-framed incentive arms. In study #2, participants were randomized to control, loss-framed incentive, normative feedback or loss-framed incentive plus normative feedback arms.

**Results:** In study #1, 1,514 nights of data were captured (69%) and sleep duration during the intervention was higher by an average of 21 (95% CI: -8, 51) and 34 (95% CI: 7, 61) minutes per night for the gain-framed and loss-framed arms, respectively, compared to controls. In study #2, 2,689 nights of data were captured (81%), with no major differences in average sleep duration between the control and the loss-framed or normative feedback arms.

**Conclusion:** We have developed and deployed a mobile health platform that can capture sleep data and remotely communicate with families. Promising candidate intervention components will be further investigated under the *optimization phase* of the MOST framework.

## Introduction

Sleep is essential for health, but youth are increasingly spending less time sleeping^1-3^. Indeed, only 47% of middle school students achieve sufficient sleep on school nights and this drops to 35% among high school students in the US^4,5^. Delaying school start times is a critical strategy that can help prevent insufficient sleep^6-12^, but this does not fully resolve the problem^10^. In addition, school sleep education programs have had limited effectiveness^13-19^. Thus, sleep promotion strategies delivered outside of the school setting must be considered.

Children are regularly seen in pediatric primary care, making this a promising setting for treating insufficient sleep in childhood, but pediatricians do not yet have effective tools for doing so^20-22^. The time needed to deliver sleep promoting behavioral treatment during brief visits, along with poor adherence to treatment, are notable barriers^20^. Interventions with behavioral sleep promotion components, and behavioral economic adherence components, that can be remotely delivered using mobile health platforms could overcome these barriers and allow for broader dissemination to reach large numbers of youth.

We are engineering a mobile health platform for pediatricians to treat insufficient sleep. This work is guided by the Multiphase Optimization Strategy (MOST) framework that includes three phases^23^: 1) the *preparation phase*, to conceptualize and initiate the testing of intervention components; 2) the *optimization phase*, to determine optimal component settings; and 3) the *evaluation phase*, to determine if the optimized intervention package is effective^23^. We have completed two preparation phase studies. Our objective is to demonstrate feasibility and present findings from two preparatory studies that included behavioral sleep promotion components and behavioral economic components, in a mobile health platform, towards promoting longer sleep duration in children.

Inspired by the effectiveness of financial incentives to promote physical activity in adults and glucose monitoring in youth with type 1 diabetes^24,25^, in study #1 we explored if providing financial incentives for achieving or exceeding sleep goals helped children to increase their sleep duration. We specifically explored the effectiveness of gain-framed and loss-framed incentives. Based on Prospect Theory, the loss-framed structure should be more motivational because of loss aversion^26^. In Study #2, we explored if providing normative feedback, with and without a loss-framed financial incentive, in relation to achieving or exceeding sleep goals could encourage children to increase their sleep duration. The normative feedback approach was inspired by a team-based approach that effectively increased physical activity in adults^27^. We created cohorts of three participants and each participant received a weekly report that informed them of their rank compared to the other cohort members with respect to the number of times they achieved or exceeded their sleep goal. Here were we present findings from both preparation phase studies.

## Methods

### Participants

Participants were enrolled in 2017 for study #1 and in 2018 for study #2, from the southeastern PA and southern NJ regions. For both studies children aged 10-12 years were eligible. We focused on a single developmental period as a starting point. Puberty can initiate at ages 10-12y, but onset and progression are more common in older children and their inclusion would have made data interpretation more challenging at this initial research stage. Participants had to have access to a tablet or smartphone to use a mobile application to transmit their sleep data remotely. Finally, participants had to typically spend 8-9 hours in bed as reported by a parent/guardian for study #1, and 7-8 hours in bed as reported by a parent/guardian in study #2. The exclusion criteria were: diagnosed with any clinical sleep disorder, syndromic obesity, diagnosed psychiatric disorder, diagnosed eating disorder, diagnosed musculoskeletal or neurological disorder that impact movement and activity, and use of medications known to affect sleep and/or body weight. All sleep data were collected during the school term, with participants starting and ending the study during a single semester. The Children’s Hospital of Philadelphia’s (CHOP) Institutional Review Board approved both studies (study #1: 17-014078; study #2: 17-014700) and both were registered at clinicaltrials.gov (study #1: NCT03263338; study #2: NCT03426644).

### Recruitment

CHOP’s Recruitment Enhancement Core compiled lists of potentially eligible children by pre-screening electronic health records of children aged 10-12y who had previously received care at CHOP. Families were invited by email to participate. Interested families opted in and were directed to a mobile health platform – called Way to Health^28^ - to complete a screening survey. If eligible, the family was invited to the baseline study visit: informed consent/assent, survey completion, height and weight measurements, and received a sleep tracker. Post-intervention, families were invited for a follow-up visit.

### Remote Data Collection and Communication

In both studies, the Fitbit Flex 2 was used to measure sleep duration in the home setting. The sleep data were captured by the Way to Health platform from the Fitbit mobile application using an application programming interface system. This approach required participants to sync their Fitbit application every day. The Fitbit Flex 2 was used for the following reasons: 1) wireless data streaming capabilities; 2) battery could be recharged; 3) Fitbits have been validated against polysomnography in children^29,30^; and 4) easy to use and desirable to wear.

The Way to Health platform was programmed to send automated messages to families in both studies. During the run-in periods, introductory messages were sent to orient families to the study. During the intervention periods, general sleep information messages were sent to provide general knowledge and to minimally engage the control groups. Goal achievement messages and, if applicable, related financial incentive and normative feedback messages, were sent during the intervention periods. Study specific methods are described below and key study similarities and differences are listed in Supplementary Table 1.

**Table 1.** Baseline characteristics of each study sample

|  | Study #1 |  |  |  | Study #2 |  |  |  |  |
| --- | --- | --- | --- | --- | --- | --- | --- | --- | --- |
|  | Overall<br>(N=30) | Control<br>(N=11) | GF<br>(N=9) | LF<br>(N=10) | Overall<br>(N=43) | Control<br>(N=10) | LF<br>(N=11) | NFB<br>(N=11) | LF + NFB<br>(N=11) |
| Age, mean (SD), years | 11.4 (0.8) | 11.7 (0.9) | 11.3 (0.9) | 11.2 (0.7) | 11.3 (0.8) | 11.1 (0.7) | 11.2 (0.6) | 11.6 (0.7) | 11.4 (0.9) |
| Female, N (%) | 15 (50.0) | 8 (72.7) | 2 (22.2) | 5 (50.0) | 23 (53.5) | 7 (70.0) | 3 (27.3) | 10 (90.9) | 3 (27.3) |
| Asian: Non-Hispanic, N (%) | 1 (3.3) | 1 (9.1) | - | - | 6 (14.0) | 1 (10.0) | 1 (9.1) | 1 (9.1) | 3 (27.3) |
| Asian: Hispanic, N (%) | - | - | - | - | - | - | - | - | - |
| Black: Non-Hispanic, N (%) | 8 (26.7) | 3 (27.3) | 2 (22.2) | 3 (30.0) | 9 (20.9) | 5 (50.0) | 3 (27.3) | 1 (9.1) | - |
| Black: Hispanic, N (%) | 3 (10.0) | 2 (18.2) | - | 1 (10.0) | 3 (7.0) | - | - | 1 (9.1) | 2 (18.2) |
| White: Non-Hispanic, N (%) | 17 (56.7) | 4 (36.4) | 7 (77.8) | 6 (60.0) | 23 (53.5) | 4 (40.0) | 7 (63.6) | 6 (54.6) | 6 (54.6) |
| White: Hispanic, N (%) | 1 (3.3) | 1 (9.1) | - | - | 2 (4.7) | - | - | 2 (18.2) | - |
| \$20-\$29K, N (%) | 1 (3.3) | - | - | 1 (10.0) | - | - | - | - | - |
| \$30-\$39K, N (%) | - | - | - | - | - | - | - | - | - |
| \$40-\$49K, N (%) | 3 (10.0) | 2 (18.2) | - | 1 (10.0) | 6 (14.0) | 1 (10.0) | 1 (9.1) | - | 4 (36.4) |
| \$50-\$59K, N (%) | 4 (13.3) | 2 (18.2) | - | 2 (20.0) | 3 (7.0) | 1 (10.0) | 1 (9.1) | - | 1 (9.1) |
| \$60-\$69K, N (%) | - | - | - | - | 1 (2.3) | - | 1 (9.1) | - | - |
| \$70-\$79K, N (%) | 3 (10.0) | 1 (9.1) | - | 2 (20.0) | 1 (2.3) | - | 1 (9.1) | - | - |
| \$80-\$89K, N (%) | - | - | - | - | 2 (4.7) | 1 (10.0) | 1 (9.1) | - | - |
| \$90-\$99K, N (%) | 5 (16.7) | 1 (9.1) | 3 (33.3) | 1 (10.0) | 2 (4.7) | 1 (10.0) | - | - | 1 (9.1) |
| ≥\$100K, N (%) | 13 (43.3) | 5 (45.5) | 6 (66.7) | 2 (20.0) | 27 (62.8) | 6 (60.0) | 6 (54.6) | 10 (90.9) | 5 (45.5) |
| Don't know/refuse, N (%) | 1 (3.3) | - | - | 1 (10.0) | 1 (2.3) | - | - | 1 (9.1) | - |
| BMI, mean (SD), kg/m<sup>2</sup> | 18.4 (3.2) | 17.9 (3.0) | 18.1 (3.0) | 19.3 (3.9) | 19.9 (4.2) | 19.7 (2.4) | 20.3 (2.5) | 20.1 (5.5) | 19.4 (5.6) |
| - Thin, N (%) | 3 (10.0) | 2 (18.2) | - | 1 (10.0) | 5 (11.6) | - | - | 2 (18.2) | 3 (27.3) |
| - Normal, N (%) | 22 (73.3) | 8 (72.7) | 7 (77.8) | 7 (70.0) | 24 (55.8) | 8 (80.0) | 7 (63.6) | 5 (45.5) | 4 (36.4) |
| - Overweight, N (%) | 3 (10.0) | 1 (9.1) | 2 (22.2) | - | 10 (23.3) | 2 (20.0) | 4 (36.4) | 2 (18.2) | 2 (18.2) |
| - Obese, N (%) | 2 (6.7) | - | - | 2 (20.0) | 4 (9.3) | - | - | 2 (18.2) | 2 (18.2) |
| Duration, mean (SD), h/night | 8.24 (0.67) | 8.16 (0.71) | 8.13 (0.77) | 8.41 (0.58) | 8.13 (0.51) | 8.16 (0.31) | 8.08 (0.56) | 8.33 (0.47) | 7.94 (0.62) |
| - School Night | 8.08 (0.87) | 7.86 (1.20) | 8.05 (0.76) | 8.36 (0.46) | 8.04 (0.58) | 8.02 (0.40) | 8.06 (0.69) | 8.15 (0.64) | 7.94 (0.60) |
| - Non-School Night | 8.43 (1.05) | 8.44 (0.85) | 8.29 (1.00) | 8.56 (1.35) | 8.24 (0.86) | 8.38 (0.93) | 8.09 (0.68) | 8.54 (0.73) | 7.97 (1.04) |
| Onset, mean (SD), hours from 00:00 | -1.66 (0.83) | -1.61 (1.23) | -1.74 (0.62) | -1.64 (0.42) | -1.20 (0.97) | -1.26 (1.40) | -1.17 (0.66) | -1.31 (0.85) | -1.06 (0.99) |
| - School Night | -1.92 (0.99) | -1.82 (1.55) | -1.95 (0.61) | -2.00 (0.41) | -1.39 (1.01) | -1.39 (1.44) | -1.37 (0.78) | -1.47 (0.86) | -1.31 (1.04) |
| - Non-School Night | -0.99 (1.04) | -1.17 (1.11) | -1.13 (0.77) | -0.65 (1.19) | -1.04 (1.32) | -1.89 (1.92) | -0.77 (0.87) | -1.08 (0.82) | -0.50 (1.19) |
| Offset, mean (SD), hours from 00:00 | 7.10 (0.70) | 7.18 (0.87) | 6.84 (0.76) | 7.25 (0.38) | 7.47 (0.92) | 7.47 (1.35) | 7.54 (0.96) | 7.50 (0.63) | 7.37 (0.76) |
| - School Night | 6.66 (0.56) | 6.57 (0.50) | 6.57 (0.72) | 6.83 (0.48) | 7.20 (1.03) | 7.20 (1.46) | 7.31 (1.16) | 7.14 (0.69) | 7.14 (0.80) |
| - Non-School Night | 8.04 (1.01) | 8.10 (1.15) | 7.54 (1.11) | 8.42 (0.54) | 7.74 (1.46) | 7.03 (2.49) | 7.98 (0.71) | 7.95 (0.94) | 7.96 (1.13) |
| Goal: 9.5 hours, N (%) | 28 (93.3) | 9 (81.8) | 9 (100) | 10 (100) | - | - | - | - | - |
| Goal: 10.0 hours, N (%) | 1 (3.3) | 1 (9.1) | - | - | - | - | - | - | - |
| Goal: 10.5 hours, N (%) | 1 (3.3) | 1 (9.1) | - | - | - | - | - | - | - |
| Initial Goal, mean (SD), h/night | - | - | - | - | 9.11 (0.70) | 9.02 (0.63) | 9.06 (0.72) | 9.44 (0.37) | 8.91 (0.93) |
| Ramp-up Goal, mean (SD), h/night | - | - | - | - | 9.33 (0.68) | 9.27 (0.63) | 9.31 (0.72) | 9.69 (0.37) | 9.01 (0.85) |

### Study #1: Run-in, Intervention and Follow-up Periods

A 1-week run-in period was used to measure baseline sleep patterns, after which parents selected one of three time in bed goals for their child: 9.5 hours, 10.0 hours or 10.5 hours per night. Based on the compromise effect, which predicts that the middle option is most likely to be selected and not the lower or higher extremes^31^, we expected that parents would be more likely to choose the 10.0 hours per night goal option. Participants were randomized to one of three study arms for 50 days, in a simple manner using a random number generator within the Way to Health platform: control, gain-framed incentive or loss-framed incentive. Blinding was not used.

During the intervention period parents of children assigned to the gain-framed arm were provided with a virtual account with $0 and could gain $1 each night their time in bed goal was achieved or exceeded. Parents of children assigned to the loss-framed arm were additionally provided with a virtual account with a $50 endowment and $1 was deducted each night the time in bed goal was not achieved. Based on Prospect Theory, the loss-framed structure should be more motivational because of loss aversion^26^. Parents were informed of their virtual account balance daily and payments were dispensed at the end of the intervention period. The incentives were directed at parents to engage them in the study. A two-week follow-up period was used to measure sleep duration post-intervention.

### Study #2: Run-in, Intervention and Follow-up Periods

A 2-week run-in period was used in study #2, with school night data (Sun-Thurs nights) from the second week used to measure baseline sleep patterns. The first week was discarded to help avoid any upward bias (i.e., increased sleep duration upon initially receiving the sleep tracker) and participants were excluded if their average time in bed was >9 hours on school nights during the second run-in week. Participants were randomized to a study arm for the 7-week intervention period, using block randomization and block sizes of four and eight. The randomization process was automated within the Way to Health platform. A time in bed goal that was 45 minutes per night above their baseline level was provided to each participant to achieve on school nights. Time in bed goals for school nights were increased by an additional 15 minutes per night at the mid-point of the intervention period. School nights were the focus because insufficient sleep is more prevalent on school nights^4,5^. Blinding was not used.

Participants were randomized to one of four study arms: control, loss-framed incentive, normative feedback, or loss-framed incentive and normative feedback combined. All participants started the study on the first Sunday following the end of their run-in period. Cohorts of three participants were manually created for those randomized to an arm with normative feedback. Participants assigned to an arm with normative feedback received daily notifications if they achieved their goal and received a report each Sunday informing them of their weekly goal tally and how they ranked compared to the other members. The parents of participants assigned to an arm with the loss-framed incentive were provided a virtual account with a $70 endowment and $2 was deducted each school night the time in bed goal was not achieved. Parents were informed of their virtual account balance daily and payments were dispensed at the end of the intervention period. A two-week follow-up period was used to measure sleep duration post-intervention.

### Qualitative Data for Study #2

Upon completion of study #2, families were invited to complete an optional semi-structured, audio-recorded telephone interview to solicit separate feedback from caregivers and children. A grounded theory approach^32^ was used to analyze qualitative data loaded into NVivo software (QSR International, Burlingame, MA). To develop the codebook, three study team members independently reviewed four interview transcripts (2 caregiver; 2 child). The generated codes were compared and the initial codebook developed was applied to four additional transcripts.

The codebook was refined and any disagreements were resolved via group consensus. The final, stable codebook was applied to the remaining transcripts. Twenty percent of the interview transcripts were double-coded for reliability purposes; the weighted kappa of 0.81 indicated strong reliability. Example interview questions and the codebook are provided in Appendix 1.

### Statistical Analysis for Quantitative Data

The same statistical approaches were applied separately in each study. Descriptive statistics are presented overall and by study arms using means and standard deviations for continuous variables, and frequencies and percentages for categorical variables. Mixed effect linear models described nights of sleep data acquired by study week, using random intercepts and slopes, and an unstructured covariance structure. Statistical interactions between study week and the following variables were tested: study arm, sex, race and self-reported household income. This was to determine if data acquirement differed by study arms and demographics.

To determine if nighttime sleep duration (hours per night) changed over the study period in the intervention groups, compared to the control arm, mixed effect linear models, with random intercepts and slopes, and an unstructured covariance structure, were used. The fixed model components included study arm assignment, time in weeks, a study arm by time in weeks interaction, and the following covariates: sex, race and household income. In study #1, weeknight status (school or non-school night) was an additional covariate. In study #2, the analyses were restricted to school nights. The mixed effect modeling was repeated with time in bed (hours per night) as an outcome. Time in bed and sleep duration were highly correlated (r=0.96 in study #1 and r=0.97 in study #2). Finally, to determine if the probability of sleeping ≥9 hours per night changed over the study period, compared to the control arm, mixed effect logistic regression was used with random intercepts and slopes, and an unstructured covariance structure. The fixed effect components for the logistic model were the same as described for the linear model. All quantitative analyses were performed using Stata version 14.2 (StataCorp, College Station, TX).

## Results

### Recruitment and Baseline Characteristics

Study #1 was completed during the fall school semester in 2017. A total of 147 Way to Health accounts were created (Figure 1). Of these, 76 parents/guardians completed the screening questionnaire; 37 were eligible and scheduled a study visit; 31 attended the study visit; 30 families selected a sleep goal and were randomized to a study arm; 22 returned for the post-intervention visit. The mean age was 11.4y and 50% were female; 60% were White, 37% were Black, and 3% were Asian; most (60%) lived in households with incomes ≥$90,000 (Table 1). Average sleep duration was 8.24 hours per night, with average time of onset and offset being 22:20 and 07:06 (Table 1). The time in bed goal of 9.5 hours per night was selected by 28 out of the 30 participants (Table 1).

**Figure 1.**
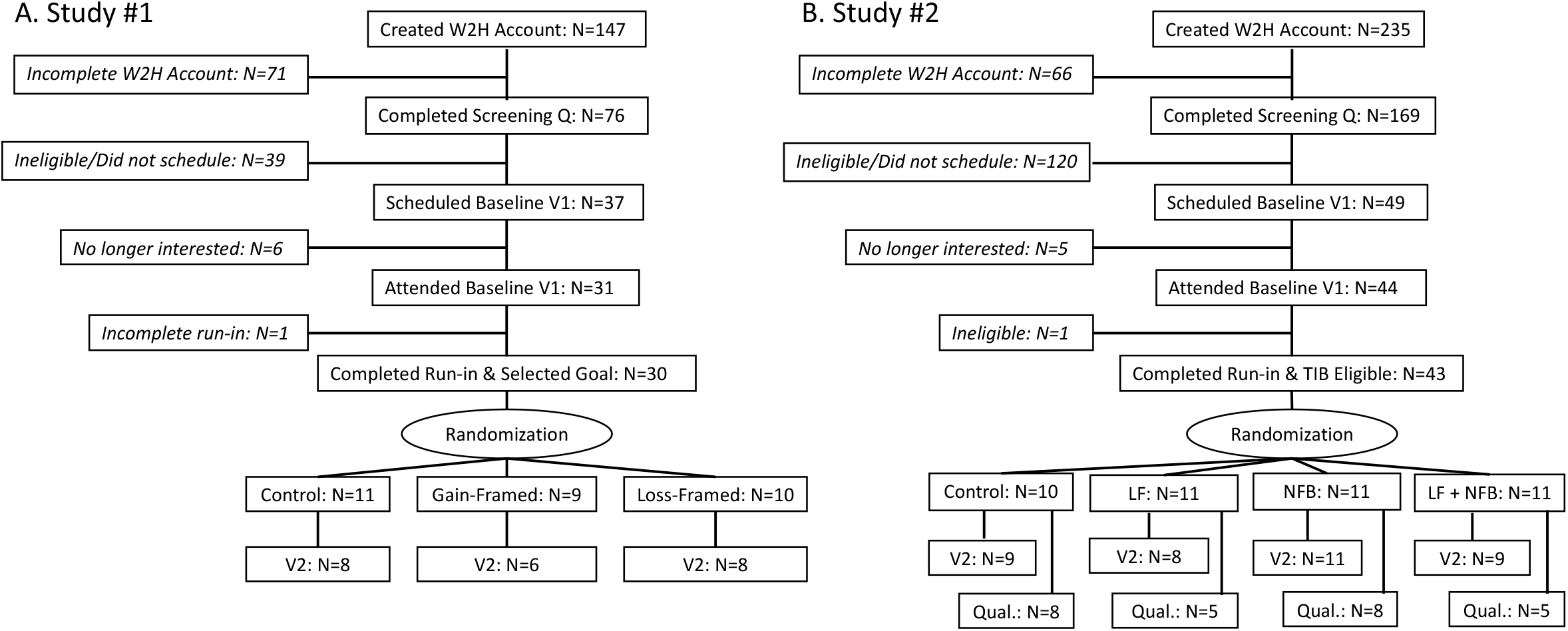
Consort diagrams for study #1 (panel A) and study #2 (panel B). Abbreviations: W2H, Way to Health platform; Q, questionnaire; V1, study visit number 1; V2, study visit number 2; TIB, time in bed; LF, loss framed incentive; NFB, normative feedback; Qual, qualitative data collection.

Study #2 was completed during the spring and fall school semesters in 2018. A total of 235 Way to Health accounts were created (Figure 1). Of these, 169 parents/guardians completed the screening questionnaire; 49 were eligible and scheduled a study visit; 44 attended the study visit; 43 were randomized to a study arm; 37 returned for the post-intervention visit; 26 families completed the optional phone interview (Figure 1). The mean age was 11.3y and 54% were female; 58% were White, 28% were Black, and 14% were Asian; most (63%) lived in households with incomes ≥$100,000 (Table 1). Average sleep duration was 8.13 hours per night, with average time of onset and offset being 22:48 and 07:28, respectively (Table 1). The average initial time in bed goal was 9.11 hours per night (Table 1).

### Sleep Data Acquisition

Overall, 1,514 nights of sleep data were collected in study #1 representing 68.8% data acquisition. On average 6.6 nights per week, 5.0 nights per week, and 3.5 nights per week were acquired during the run-in, intervention and follow-up periods, respectively (Figure 2). Data acquisition was similar for each study arm, both sexes and household income groups (Figure 2). Compared to White participants, data acquisition among Black participants was 2.2 (95% CI: 0.8, 3.7) nights per week lower during the intervention period (Figure 2).

**Figure 2.**
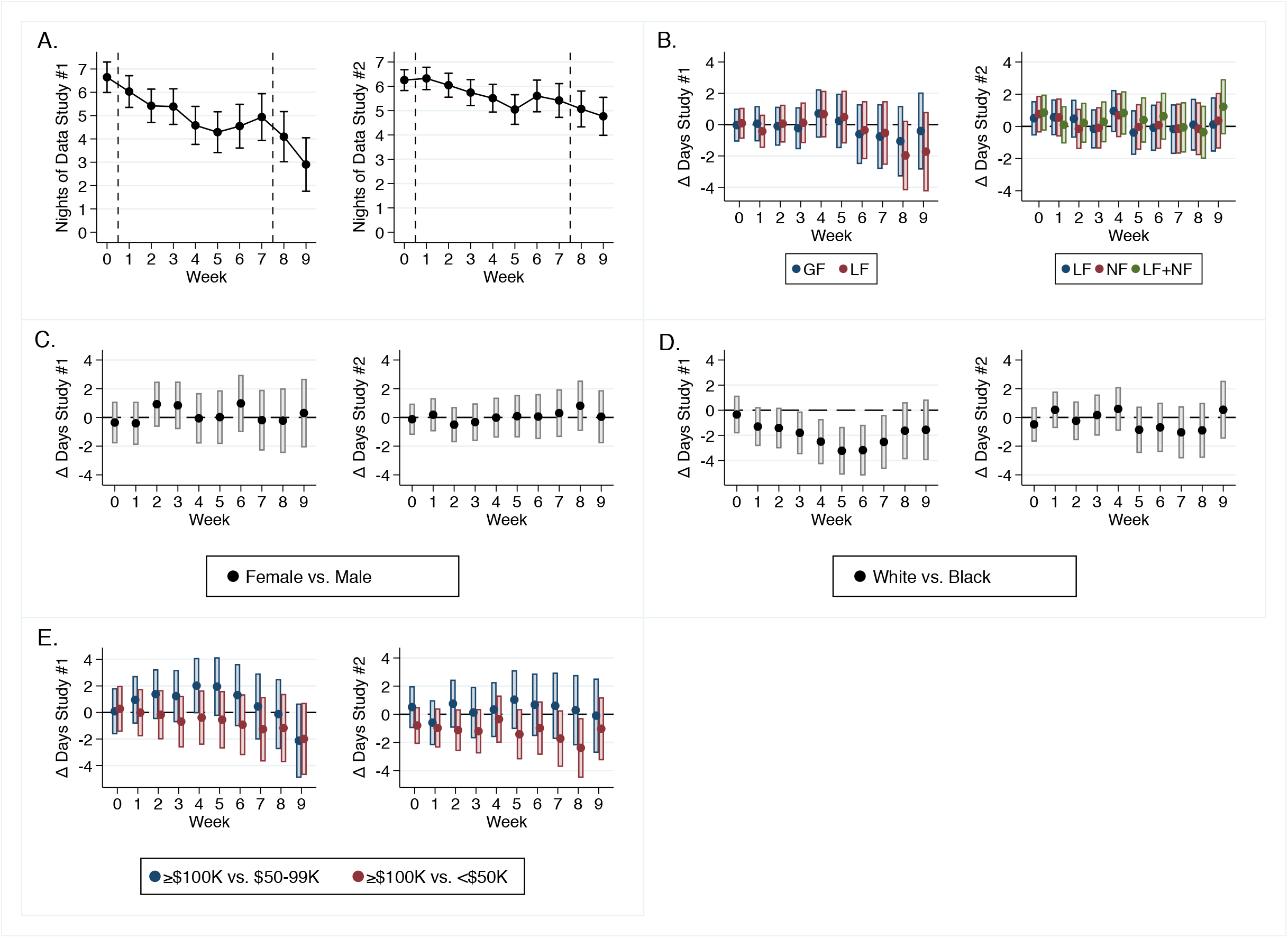
Average nights of sleep data acquired in each study, by study week, with 95% confidence intervals. Panel A. Average acquirement for study #1 (left) and study #2 (right). Panel B. Difference in average nights acquired by study arm, for study #1 (left) and study #2 (right). Abbreviations: C, control arm; GF, gain-framed arm; LF, loss-framed arm; and NF, normative feedback arm. Panel C. Difference in average nights acquired by sex for study #1 (left) and study #2 (right). Panel D. Difference in average nights acquired by race for study #1 (left) and study #2 (right). Due to the small number of participants the Asian race category is not presented. Panel E. Difference in average nights acquired by household income for study #1 (left) and study #2 (right).

In study #2, 2,689 nights of sleep data were collected representing 81.2% data acquisition. On average 6.3 nights per week, 5.7 nights per week, and 4.9 nights per week were acquired during the run-in, intervention and follow-up periods, respectively (Figure 2). Data acquisition was similar for each study arm, both sexes, and race and household income groups (Figure 2).

### Changes in Sleep by Study Arm Assignment

In study #1, sleep duration during the overall 50-day intervention period was higher by an average of 0.36 hours per night (95% CI: -0.13, 0.85) and 0.57 hours per night (95% CI: 0.11, 1.02) for the gain-framed and loss-framed arms, respectively, compared to the control arm (Figure 3). During the follow-up period, sleep duration was higher by an average of 0.55 hours per night (95% CI: -0.01, 1.11) and 0.34 hours per night (95% CI: -0.20, 0.89) for the gain-framed and loss-framed arms, respectively, compared to the control arm (Figure 3). Similar findings were observed when using time in bed as the outcome (Supplementary Figure 1). The probability of sleeping ≥9 hours per night during the overall 50-day intervention period was higher by an average of 10.0 percentage points (95% CI: -5.0, 25.4) and 18.0 percentage points (95% CI: 3.3, 32.6) for the gain-framed and loss-framed arms, respectively, compared to the control arm (Figure 4). And was also higher for the gain-framed and loss-framed arms during the follow-up period by an average of 12.1 percentage points (95% CI: -7.0, 31.0) and 5.8 percentage points (95% CI: -12.2, 23.8), respectively, compared to the control arm (Figure 3).

**Figure 3.**
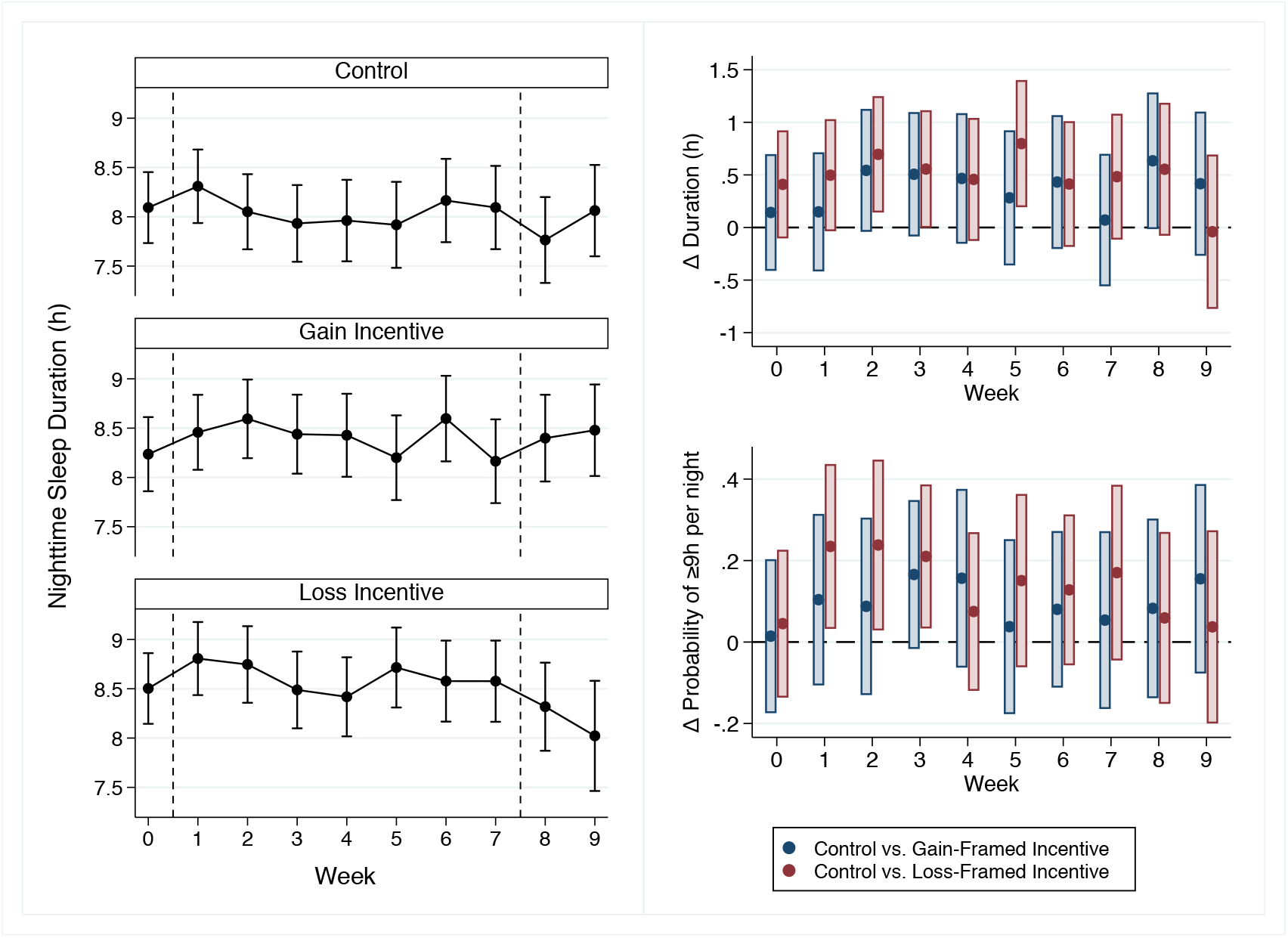
Changes in nighttime sleep duration for study #1. The left column presents averages and 95% confidence intervals for sleep duration (hours per night) by study arm and study week. The column on the right presents the difference in sleep duration and the probability of sleeping ≥9 hours per night by study week and study arms, relative to the control arm.

**Figure 4.**
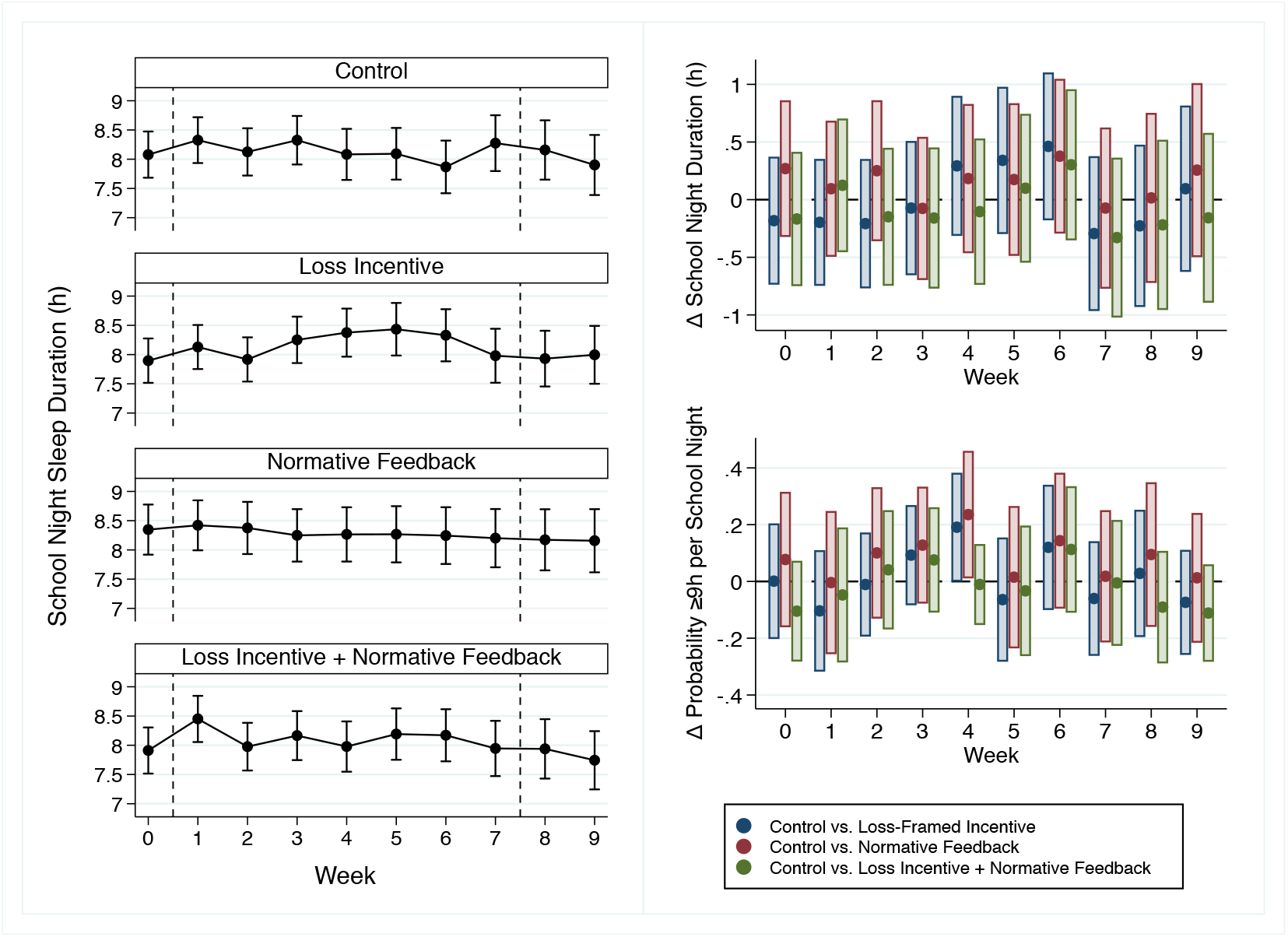
Changes in nighttime sleep duration for study #2. The left column presents averages and 95% confidence intervals for sleep duration (hours per school night) by study arm and study week. The column on the right presents the difference in school night sleep duration and the probability of sleeping ≥9 hours per school night by study week and study arms, relative to the control arm.

In study #2, two out of the eight cohorts assigned to a normative feedback arm included two members (Supplementary Table 2). Delayed intervention starts occurred in five of the cohorts, with three participants having delays of 1-2 weeks and two participants having a delay of 26 weeks (Supplementary Table 2); meaning, members of these cohorts were not always active on study at the same time to provide data for the weekly rankings.

In study #2, school night sleep duration during the overall 7-week intervention period differed on average by 0.23 hours per night (95% CI: -0.37, 0.83), 0.11 hours per night (95% CI: -0.41, 0.64), and 0.12 hours per night (95% CI: -0.52, 0.76) for the loss-framed arm, normative feedback arm, and the combined arm, respectively, compared to the control arm (Figure 4). During the follow-up period, school night sleep duration differences with the control arm averaged -0.22 hours per night (95% CI: -0.81, 0.37), -0.06 hours per night (95% CI: -0.58, 0.45), and -0.23 hours per night (95% CI: -0.86, 0.41) for loss-framed arm, normative feedback arm, and the combined arm, respectively (Figure 4). Similar findings were observed when using time in bed as the outcome (Supplementary Figure 2). The probability of sleeping ≥9 hours per school night during the overall 7-week intervention period differed on average by 8.1 percentage points (95% CI: -14.6, 30.8), 9.0 percentage points (95% CI: -7.4, 25.4), and 5.7 percentage points (95% CI: -13.9, 25.3) for the loss-framed arm, normative feedback arm, and the combined arm, respectively, compared to the control arm (Figure 4). And for the follow-up period, the probability of sleeping ≥9 hours per school night differed on average by -10.0 percentage points (95% CI: -27.3, 7.2), 2.4 percentage points (95% CI: -12.3, 17.1), and -9.6 percentage points (95% CI: -23.8, 4.6) for the loss-framed arm, normative feedback arm, and the combined arm, respectively, compared to the control arm (Figure 4).

### Family Feedback for Study #2

Supplementary Table 3 provides descriptive statistics for the subsample that completed the optional telephone interview. Key themes and quotations from child and caregivers are shown in Appendix A, Supplementary Tables 4 and 5. Qualitative data revealed a number of intervention facilitators, including: *enhanced sleep focus and knowledge* (e.g., increased awareness of sleep routines and importance of sleep) and *benefits to child health and wellbeing* outside of sleep (e.g., use of the Fitbit to encourage exercise). Interestingly, *electronics rules and norms* emerged as an intervention facilitator in cases where families had pre-existing evening electronics usage rules, but as an intervention barrier for families who reported difficulties limiting device usage consistently, especially when needed for homework. Salient intervention barriers included: *beliefs about electronics usage* (e.g., electronics devices are “addicting” and therefore difficult to limit), *competing academic demands and schedules* and *competing extracurricular activities* (e.g., homework, school start times, and late evening activities), *family context and schedules* (e.g., sibling activities and caregiver work schedules limit consistent sleep routines), and *long-term sustainability* (e.g., challenge of maintaining optimal sleep schedule and duration after study ended). Of note, children and caregivers also indicated a *desire for intervention personalization* in both intervention content (e.g., sleep information and advice) and style (e.g., graphics and photos).

## Discussion

We have developed and deployed a mobile health platform that can remotely capture sleep data from children. This platform can also remotely communicate behavior change strategies toward promoting sleep duration to families in the home setting. For example, in study #1 we used a guideline-based sleep goal paired with a loss-framed incentive directed at parents; compared to the control arm, the loss-framed incentive arm increased sleep duration by an average of 34 minutes per night and increased the probability of sufficient sleep (≥9 hours per night) by an average of 18 percentage points. However, this promising loss-framed incentive finding was not replicated in study #2. Guided by the MOST framework, these studies represent essential preparation phase research needed to inform a future optimization trial as we work towards engineering a mobile health platform for pediatricians to support families at home for the promotion of childhood sleep duration.

Nights of data captured improved from 69% to 81% as we progressed from study #1 to study #2. We acquired fewer nights of data in study #1 from Black versus White participants, but this protocol disparity was not observed in study #2, indicating that the mobile health platform in development is feasible and can engage a diverse scope of participants. The adherence improvements are likely the consequence of a more rigorous process to actively monitor adherence to the study protocol and the ease of doing so with the structured study start in study #2 (i.e., intervention periods started on the first Sunday following the end of the run-in period).

Setting a sleep goal is a key intervention component for sleep extension^33-35^. In study #1, we offered three time in bed goals, all based around the sleep duration guideline. When presented with three choices, the compromise effect predicts that the middle option is most likely to be selected and not the lower or higher extremes^31^. However, 28 out of 30 participants selected the lowest option indicating that the goals we offered may have been too challenging. Self-efficacy theory predicts that children will be more motivated to increase their sleep duration if they believe that they can attain the goal^36^. In study #2, we used a personalized approach that set time in bed goals to be 45 minutes per night higher than baseline, with an additional 15 minute per night ramp up mid-study. This approach may be more conducive to enhancing self-efficacy, but assumes that the baseline measurement correctly captures typical sleep duration and that the initial and ramp up goals that we set are realistic and achievable. In a future optimization trial, we will further investigate goal component settings. Optimizing the sleep goal component is especially important when other intervention components are tied to achieving or exceeding the goal, such as with financial incentives and normative feedback.

Loss-framed financial incentives have been used in prior studies to effectively change behavior^24,25^. Based on Prospect Theory, the loss-framed structure should be more motivational than the gain-framed structure because of loss aversion^26^. In study #1, preliminary findings support the application of a parent-directed loss-framed financial incentive to encourage sleep promotion in children, but this was not replicated in study #2. It is important to highlight that time in bed goals tied to the incentives (guideline-based for all nights versus personalized on school nights) and incentive values ($1 versus $2) differed between the two studies. More research is needed to determine the most optimal application of the loss-framed incentive for sleep promotion. In both studies the money was dispensed at the end of the intervention period and dispensing more frequently may be more effective. We directed the incentives at parents and effectiveness may be improved if directed at children. Finally, the longer-term implications of incentives on establishing lasting changes in behavior to promote sleep needs to be investigated.

In study #2, the normative feedback approach used was not associated with meaningful changes in sleep duration, with or without a loss-framed incentive. We failed to correctly align the start times for five out of the eight normative feedback cohorts, meaning that members of those cohorts were not always active on study at the same time to provide data for the weekly rankings. These protocol deviations may have impacted effectiveness. Further, cohorts were created based on the order of recruitment and membership was not contingent on having any similar (or dissimilar) characteristics. The cohort approach was adopted to facilitate feedback because we do not yet have extensive reference data from which to make normative comparisons. It would be of interest to investigate this further after more data are generated and representative reference data become available.

In study #2, qualitative interview data provided important information about intervention barriers and facilitators. Barriers to sleep extension were primarily related to competing academic and extracurricular demands, as well as sibling activities and caregiver work schedules. Families reported challenges around creating consistent rules related to electronic device usage. In line with findings from a recent qualitative study with adolescents^37^, children described feeling “addicted” to electronic devices. Of note, child participants desired more personalized messaging to target their specific sleep habits, while caregivers suggested enhancing the style of the message content (e.g., photos). Based on this feedback, a future candidate component could focus on delivering more personalized, evidence-based guidance to overcome barriers to changing sleep patterns, particularly around electronics rules, norms, and beliefs.

Both studies included limitations. We disproportionately enrolled more White and higher income participants. There are established sex, race and income disparities in sleep health: male children (vs. female), Black children (vs. White) and children from lower income households (vs. higher income) tend to have shorter sleep duration^38-40^. The recruitment approach used in both studies used email invitations. In future studies, it will be critical to adopt additional recruitment strategies to ensure a more diverse sample. Otherwise the mobile health platform we are engineering will be less effective for the children in most need of sleep promotion treatment. We used a commercial sleep tracker and the sleep scoring algorithm used is proprietary^29,30^. As sleep tracker technology advances it will be important to consider a device that provides access to the raw data captured by the device sensors and the application of transparent and validated sleep scoring algorithm for children. In addition, participants were required to sync a mobile application so that their sleep data could be retrieved by the Way to Health platform. Removing this step would improve the flow of data collection. This could be accomplished by using a device that has built-in wi-fi to transmit the data and would negate the need for participants to have access to a smartphone or tablet computer to be eligible.

In conclusion, we have developed and deployed a mobile health platform that is capable of capturing sleep data and remotely communicating with families towards promoting longer sleep among middle-school aged children. We will further investigate promising candidate intervention components in a future optimization trial.

## Supporting information

Consort Checklist

## Data Availability

The data can be made available upon request to the corresponding author (JM). Access to the data will require a data sharing agreement with the Children's Hospital of Philadelphia.

## Acknowledgements

We thank the families for volunteering to take part in this study. We appreciate the dedication of the research staff who helped to collect and manage the data. Study #1 was supported by an NIH/NHLBI Career Development Award K01HL123612 and internal awards from the Children’s Hospital of Philadelphia (Pediatric Development Funds and the Metabolism, Nutrition and Physical Development Research Affinity Group). Study #2 was supported by an NIH/NHLBI Career Development Award K01HL123612 and in part by Grant Number UL1TR001878 from the National Center for Advancing Translational Science. The content is solely the responsibility of the authors and does not necessarily represent the official views of the National Center for Advancing Translational Science or the National Institutes of Health. Study #2 was also supported in part by the Institute for Translational Medicine and Therapeutics at the University of Pennsylvania. Ariel A. Williamson was supported by a Eunice Kennedy Shriver National Institute of Child Health and Human Development Career Development Award K23HD094905 and by the Sleep Research Society Foundation.

