## Supplementary File for "Engineering a Mobile Platform to Promote Sleep in the Pediatric Primary Care Setting"

Supplementary Table 1 – Summary of differences between the two preparation phase studies.

| | Study #1 | Study #2 |
|---|---|---|
| Data collection | 2017 | 2018 |
| School semester | Fall | Spring and Fall |
| Age | 10-12y | 10-12y |
| Sample size | 30 | 43 |
| Sleep tracker | Fitbit Flex 2 | Fitbit Flex 2 |
| Time in bed eligibility | 8-9 hours per night | 7-8 hours per night |
| Run-in | 7 days | 14 days |
| Run-in exclusion | None | Avg. time in bed >9 h on school nights |
| Time in bed goal | Guideline-based, all nights | Personalized, school nights |
| Intervention start day | First day after run-in ended | First Sunday after run-in ended |
| Intervention length | 50 days | 49 days |
| Study arms | Control<br>Gain-framed incentive ($1 value)<br>Loss-framed incentive ($1 value) | Control<br>Loss-framed incentive ($2 value)<br>Normative feedback<br>Loss-framed ($2 value) + normative feedback |
| Follow-up | 14 days | 14 days |

Supplementary Table 2 – Cohort details for the normative feedback arms in study #2

| | | | Start time difference (weeks) with Member #1... | |
|---|---|---|---|---|
| Cohort | Study Arm | Total # Members | Member #2 | Member #3 |
| A | Normative Feedback | 3 | 0 | 0 |
| B | Normative Feedback | 3 | 0 | 26 |
| C | Normative Feedback | 3 | 0 | 0 |
| D | Normative Feedback | 2 | 2 | - |
| E | Loss-Framed Incentive + Normative Feedback | 3 | 0 | 1 |
| F | Loss-Framed Incentive + Normative Feedback | 3 | 0 | 26 |
| G | Loss-Framed Incentive + Normative Feedback | 3 | 0 | 0 |
| H | Loss-Framed Incentive + Normative Feedback | 2 | 1 | - |

Supplementary Table 3 – Description of the subsample who completed a phone interview in study #2

| | Overall (N=26) | Control (N=8) | LF (N=5) | NFB (N=8) | LF + NFB (N=5) |
|---|---|---|---|---|---|
| Age, mean (SD), years | 11.3 (0.7) | 11.1 (0.7) | 11.1 (0.6) | 11.6 (0.7) | 11.3 (0.8) |
| Female, N (%) | 17 (65.4) | 5 (62.5) | 3 (60.0) | 7 (87.5) | 2 (40.0) |
| Asian: Non-Hispanic, N (%) | 3 (11.5) | 1 (12.5) | - | 1 (12.5) | 1 (20.0) |
| Asian: Hispanic, N (%) | - | - | - | - | - |
| Black: Non-Hispanic, N (%) | 7 (29.6) | 4 (50.0) | 2 (40.0) | 1 (12.5) | - |
| Black: Hispanic, N (%) | 2 (7.7) | - | - | 1 (12.5) | 1 (20.0) |
| White: Non-Hispanic, N (%) | 13 (50.0) | 3 (37.5) | 3 (60.0) | 4 (50.0) | 3 (60.0) |
| White: Hispanic, N (%) | 1 (3.9) | - | - | 1 (12.5) | - |
| $20-$29K, N (%) | - | - | - | - | - |
| $30-$39K, N (%) | - | - | - | - | - |
| $40-$49K, N (%) | 2 (7.7) | - | 1 (20.0) | - | 1 (20.0) |
| $50-$59K, N (%) | 1 (3.9) | 1 (12.5) | - | - | - |
| $60-$69K, N (%) | - | - | - | - | - |
| $70-$79K, N (%) | 1 (3.9) | - | 1 (20.0) | - | - |
| $80-$89K, N (%) | - | - | - | - | - |
| $90-$99K, N (%) | 1 (3.9) | 1 (12.5) | - | - | - |
| ≥$100K, N (%) | 20 (76.9) | 6 (75.0) | 3 (60.0) | 7 (87.5) | 4 (80.0) |
| Don’t know/refuse, N (%) | 1 (3.9) | - | - | 1 (12.5) | - |
| BMI, mean (SD), $kg/m^2$ | 19.8 (4.2) | 19.0 (2.3) | 20.5 (3.4) | 18.7 (4.6) | 22.0 (6.3) |
| - Thin, N (%) | 2 (7.7) | - | - | 2 (25.0) | - |
| - Normal, N (%) | 16 (61.5) | 7 (87.5) | 3 (60.0) | 4 (50.0) | 2 (40.0) |
| - Overweight, N (%) | 5 (19.2) | 1 (12.5) | 2 (40.0) | 1 (12.5) | 1 (20.0) |
| - Obese, N (%) | 3 (11.5) | - | - | 1 (12.5) | 2 (40.0) |
| Duration, mean (SD), h/night | 8.03 (0.55) | 8.14 (0.35) | 7.74 (0.54) | 8.34 (0.48) | 7.65 (0.69) |
| - School Night | 7.93 (0.60) | 8.00 (0.45) | 7.63 (0.64) | 8.23 (0.58) | 7.64 (0.70) |
| - Non-School Night | 8.22 (0.76) | 8.45 (0.49) | 7.99 (0.79) | 8.46 (0.84) | 7.70 (0.86) |
| Onset, mean (SD), hours from 00:00 | -1.28 (0.77) | -1.72 (0.58) | -1.01 (0.53) | -1.39 (0.88) | -0.66 (0.69) |
| - School Night | -1.44 (0.81) | -1.92 (0.47) | -1.14 (0.47) | -1.48 (0.97) | -0.90 (0.96) |
| - Non-School Night | -0.90 (0.96) | -1.19 (0.72) | -0.67 (0.91) | -1.27 (0.73) | -0.06 (0.68) |
| Offset, mean (SD), hours from 00:00 | 7.27 (0.57) | 6.99 (0.38) | 7.27 (0.44) | 7.45 (0.58) | 7.42 (0.87) |
| - School Night | 7.01 (0.66) | 6.67 (0.39) | 7.01 (0.67) | 7.25 (0.71) | 7.16 (0.86) |
| - Non-School Night | 7.85 (0.82) | 7.81 (0.64) | 7.90 (0.24) | 7.68 (0.72) | 8.17 (1.52) |

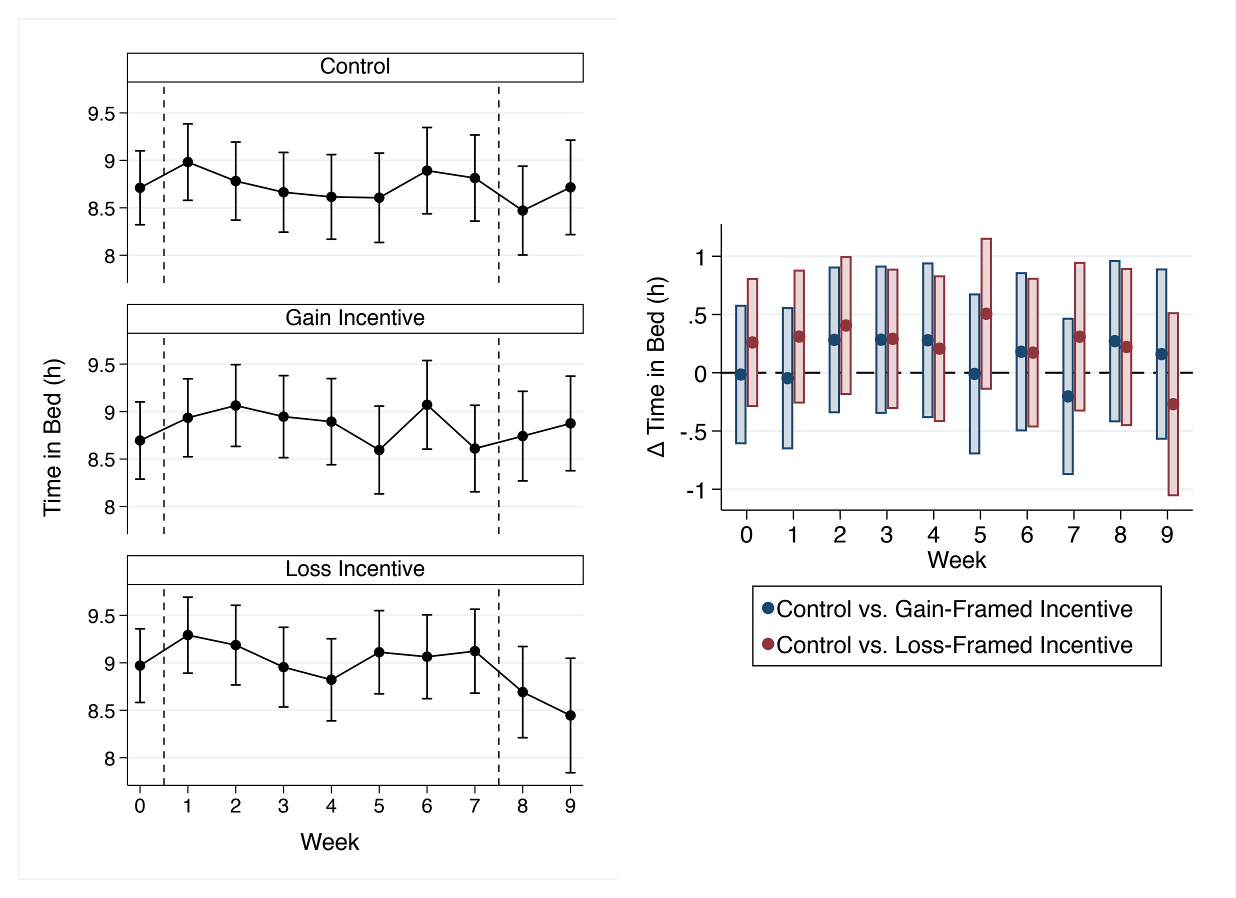

Supplementary Figure 1. Changes in time in bed for study #1. The left column presents averages and 95% confidence intervals for time in bed (hours per night) by study arm and study week. The column on the right presents the difference in time in bed by study week and study arms, relative to the control arm. The overall difference between the control arm and the gain-framed arm during the intervention period and follow-up periods were: 0.11 (95% CI: -0.41, 0.64) hours per night and 0.23 (95% CI: -0.39, 0.85) hours per night. The overall difference between the control arm and the loss-framed arm during the intervention period and follow-up periods were: 0.32 (95% CI: -0.17, 0.80) hours per night and 0.04 (95% CI: -0.56, 0.65) hours per night.

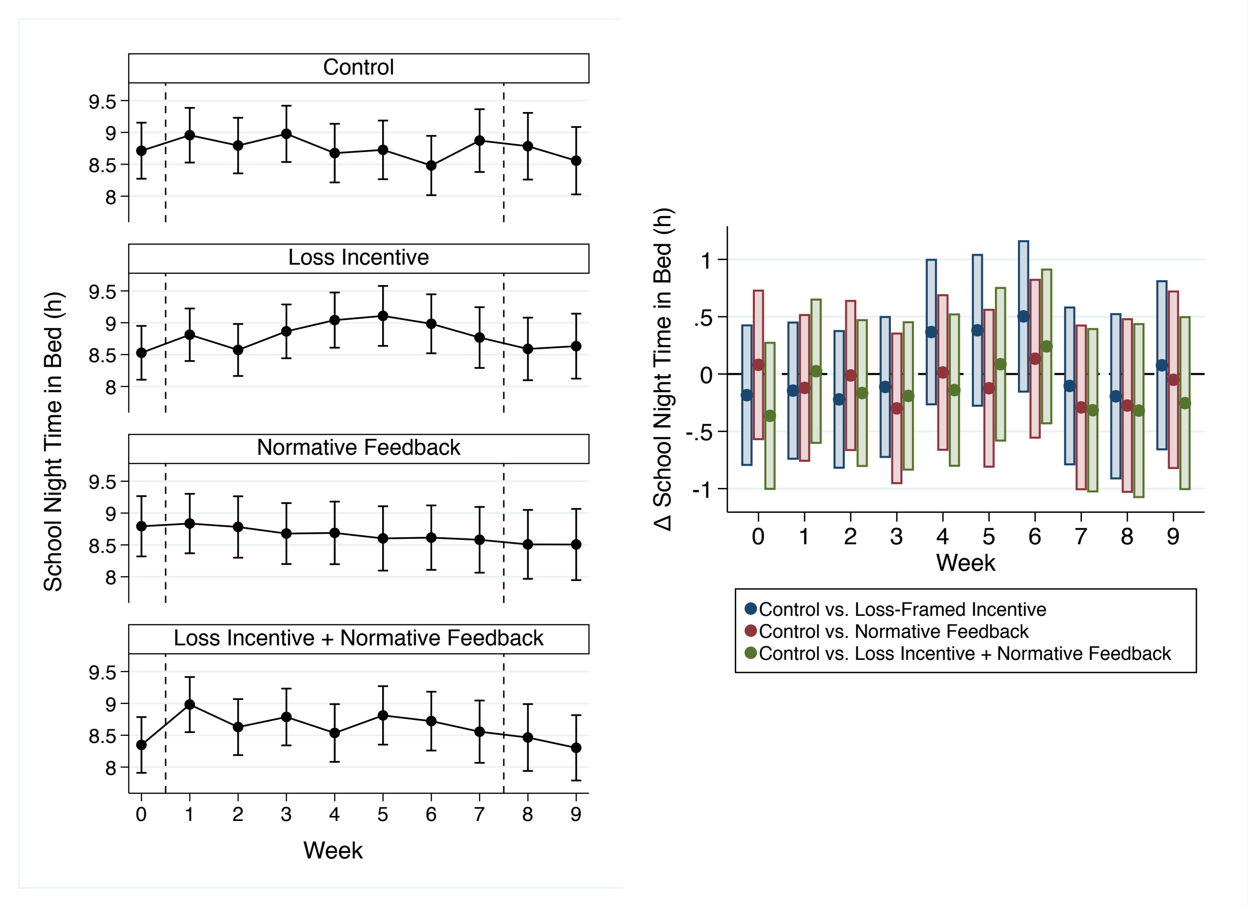

Supplementary Figure 2. Changes in time in bed for study #2. The left column presents averages and 95% confidence intervals for time in bed (hours per school night) by study arm and study week. The column on the right presents the difference in time in bed by study week and study arms, relative to the control arm. The overall difference between the control arm and the loss-framed arm during the intervention period and follow-up periods were: 0.06 (95% CI: -0.44, 0.56) hours per night and -0.06 (95% CI: -0.66, 0.54) hours per night. The overall difference between the control arm and the normative feedback arm during the intervention and follow-up periods were: -0.12 (95% CI: -0.67, 0.43) hours per night and -0.17 (95% CI: -0.82, 0.48) hours per night. The overall difference between the control arm and the combined arm during the intervention and follow-up periods were: -0.10 (95% CI: -0.64, 0.44) hours per night and -0.32 (95% CI: -0.95, 0.32) hours per night.

**Appendix A: Qualitative Data Collection Information and Codebook**

<u>Sample caregiver interview questions</u>

**Overall changes to child sleep; general barriers and facilitators:**

- During the study, did you try to make any changes to increase your child's sleep?
    - (If yes) Tell me more about this. What kinds of things did you do? Did anything get in the way? Did anything make this easier?
    - Are you still doing ____? Tell me more about that.
    - (If no) Tell me more about that decision.
- Tell me about any changes you would recommend to other parents trying to help their children get more sleep.

**Sleep-wake routine questions:**

- Part of the study was to try to get your child in bed earlier. Did you change any part of the bedtime routine to help with this?
    - (If yes) Tell me more about that, what kinds of things did you change?
    - (If no) Tell me more about that decision.
- What about following a bedtime routine?
    - Was there anything that made it hard or got in the way of following a bedtime routine?

**Sleep health behavior questions:**

- Some families have found that a regular bedtime- meaning having the same bedtime every night- helps their children get more sleep. Other families have found that this is not helpful. What do you think?
    - What kind of things get in the way of having a regular bedtime?
    - What kind of things help with having a regular bedtime?
- Did you have any rules about the use of electronics before bedtime or overnight during the sleep study? Tell me more about that. Do you have the same rules now?
    - Tell me about the weekends/Do the same rules apply?

**Study procedures:**

- Now I want to ask you about how the study was set up—things like the number of visits, the visit location, and other things. Tell me what you think about how the study was set up (whatever comes to mind).
- You and your child received messaging from us with reminders and sleep tips. What did you think about this?
    - Do you have any suggestions to make the messaging better?
- This study used a Fitbit- as a parent, what did you think about your child using the Fitbit?
    - How was using the app?
    - Do you think the Fitbit and app really represented the way your child slept?

**General:**

- Is there anything else you would like me to know about the study and what it was like for you and your family?

Sample child interview questions

**Overall changes to child sleep; general barriers and facilitators:**

- The Sleep Study that you were part of asked families to help their child get more sleep. During the study, did you try to make any changes to increase your sleep?
  - (If yes) Tell me more about this. What kinds of things did you do?
  - Did anything get in the way or make this hard? Did anything make this easier?
  - Are you still doing ____? Tell me more about that.
  - (If no) Tell me more about this. Did anything get in the way of making changes?

**Sleep-wake routine questions:**

- Part of the study was to try to get to bed earlier. Did you change anything in the routine to help with this?
  - (If yes) Tell me more about that, what kinds of things did you change?
  - (If no) Tell me more about that decision.
  - Was there anything that made it hard or got in the way of getting in to bed earlier?
- What about following a bedtime routine?
  - Was there anything that made it hard or got in the way of following a bedtime routine

**Sleep health behavior questions:**

- What do you think about setting a regular bedtime on a weeknight? By "regular" I mean a bedtime that is pretty much the same time every night.
  - What kind of things get in the way of having a regular bedtime?
  - What kind of things help with having a regular bedtime?
- Did you have any rules at home about using electronics before bedtime or overnight during the Sleep Study?
  - Tell me more about that. Do you have the same rules now?

**Study procedures:**

- Now I want to ask you about how the study was set up—things like the number of visits, the visit location, and other things. Tell me what you think about how the study was set up (whatever comes to mind).
- Do you have any suggestions to make the study better or easier to follow?
- Your parent received messaging from us with reminders and sleep tips. What did you think about this?
  - Do you have any suggestions to make the messaging better? What about sending messages to kids and to parents?
- This study used a Fitbit- how was using the Fitbit?
  - How was using the app?
  - Do you think the Fitbit and app really showed the way you actually slept?

Supplementary Table 4 - The final qualitative codebook

| Code | Sub Code | Definition |
|---|---|---|
| Enhanced Sleep Focus/Sleep Engagement | | Increased intentionality, impact on family prioritizing sleep, enhanced sleep focus (parental engagement), accountability. |
| Competing Child Activities | School Start Time | Any mention of early start time, early bus pick-up, or less flexibility in morning schedule. |
| | Homework | Mention of homework/academic obligations |
| | Extracurricular | After school activity, sport, club. Could include dance, music, theater, art, etc. |
| | Social, Work, Chores | Social or work activity (employment; hanging out with friend, etc.) or chores at home |
| Electronics Usage | Study Impact on Usage | Any reference to impact of study participation in electronics usage or new awareness of electronics usage. Include rules set as a result of the study or rules that were ignored/hard to implement. |
| | Family Rules/ Norms about Device Usage | A specific rule or norm/ habit followed by the child or family about electronic usage implemented (before, during and/or after the study period). This can include statements like "As a family we usually say no electronics on weekdays." |
| | Beliefs about Electronics Usage | Any mention of device usage and its effect on sleep or what families/kids think about devices in general ("they are distracting/bad for you/etc." without mention of sleep). |
| Study Impact on Sleep Routines and Duration | | Change in routine due to study, increased sleep due to study, any mention of study involvement affecting the child's sleep. |
| Parent-Child Communication | | Any reference to communication between child and parent during the study, how they handled study procedures, conversed about messaging. |
| Family Beliefs About Sleep | | Beliefs, opinions, or thoughts that anyone in family expresses about sleep. |
| Competing Family Demands | | Impact of siblings on sleep, bedtime or morning routines; caregiver/parent work schedules that impact sleep. |
| Long Term Sustainability | | Any reference to whether changes to sleep or healthy sleep habits have been maintained post-study participation |
| Feasibility/Acceptability | Fitbit | Any mention of ease of use of the Fitbit and app; perception of Fitbit validity/measurement of sleep |
| | Messaging | References to messaging from the app or the study team |
| | Study Procedures | Any mention of sleep journals, study visits, surveys, or communicating with staff |

| | | |
|---|---|---|
| Ancillary Benefits | | Additional benefit of study that benefitted the child or family in some positive way, outside of sleep. |
| Personalized Approach | | References to parent or child desire for more individualized approach to messaging, goal-setting, meeting Fitbit parameters, working on barriers to achieving goals, etc. |

Supplementary Table 5 - Qualitative themes and example participant quotes

| Theme | Child participant quote | Caregiver participant quote |
|---|---|---|
| Intervention facilitators | | |
| Enhanced sleep knowledge and focus | *"…um, I would say just like at nighttime before I went to bed just get everything set up for the next day so that instead of like say getting up at 6, I could get up at 630 and like my backpack would already be set up I wouldn't have to do much other than get up and get ready for school."*<br><br>*"Um it [having a regular bedtime] is helpful. Um because like um kids who like don't have those rules will go to bed really late and then they won't really have a good day the next day so it sort of like um effects how much you sleep and well it affects your day so."* | *"Um, so I think that, you know, having just some accountability, uh, definitely made us more conscious of trying to get to sleep at um, a certain time, and um, yeah, and just being more consistent and planning ahead, being just like more prepared. So I think yeah, just like having that like sense of accountability I think is what like really forced me to try to make a change."*<br><br>*"… I think that it was fun to do to just see really how much she was sleeping and to be more aware of that um you know and knowing that she should get more sleep so it was eye opening for me in that way so I appreciated that as a parent."* |
| Benefits to child health and wellbeing | *"…I mean I used it [Fitbit] for not only the sleeping thing, I just checked out what kind of fitness things I was doing. Um, I checked the steps. I did all of that. I didn't – I thought it was pretty cool because I always know – I know that Fitbit sickness that I always had secretly wanted one."*<br><br>*"Um, well, my mom and I like had competitions about who would like walk the furthest or like who got more steps and more exercise and stuff outside."* | *"…what I really liked was wearing the Fitbit raised his level of consciousness about the exercise that he's getting, and he challenged himself every day to get his 10,000 steps and even lost weight during it."*<br><br>*"He didn't wake up grumpy. He woke up and he was like he just felt better and he was better during the day and he said he never got tired at his desk because some days he admitted to me that he would like fall asleep or get kind of you know."* |
| Intervention acceptability | *"I think that is was actually a good experience like yeah because it really improved my sleeping better and my exercise and how many steps I took. I just really think that it improved me."* | *"I found it very easy to participate. I didn't think it, like, the time commitment was excessive or anything like that, and she enjoyed participating in it so, you know, that made it feel more worthwhile for me."* |

| | | |
|---|---|---|
| Beliefs about electronics usage* | *"Um, but yeah, sometimes my parents will tell me to not use it, and I won't, and, um, or I won't use it for like two hours when I usually use it, and it'll just stop - it'll never stick in the habit, though because I'm addicted to my phone."*<br><br>*"Sometimes it's good to put it away so you don't focus on it when you go to bed so you stop thinking about it but then other times it [turning off devices] just keeps you thinking about things so you can't really get to bed because there is always something you are thinking about."* | *"You know, we'd like her -- she's supposed to be off of her phone by, I think it's like 10:00, you know. But we still allow her TV to be on. She doesn't really watch it, it's just kind of like almost a white noise more than anything."*<br><br>*"I think when she uses it [cell phone] more often, she seems to have a harder time sleeping, and my thought is just because her brain is still actively going because of being on the screen and either texting or messaging people, so I think sometimes that does, um, hinder her having a more restful sleep."* |
| Intervention barriers | | |
| Electronics rules and norms | *"Um, I think that my family, we don't really limit uh, electronics, but it should be. We're going to begin to do that or else um, your kid could stay up all night playing games and being on their electronics."* | *"Well, the kids cry for it. They want it, and that's what they want to do on their free time, and you want to give them down time after a hard day's work fulfilling all their commitments, and of course that's what they want to do. It tends to be later when you get the free time, so that's a bit of a struggle. Every night, actually, it's a struggle, like, 'Time to get off.'"* |
| Family context and schedules | *"Um I have two younger sisters so they were always like screaming while I was trying to go to sleep."* | *"I think sometimes just when, like, her siblings have an activity or parents are doing other things that we're not monitoring her routine as closely to make sure that she's taking the steps to be ready."* |
| Competing academic demands and schedules | *"...But then on the weekdays you have to be most – sometimes multitasking at work and on school days it's just stressful because of all the homework and all the work that you got to turn in to other classes, especially when like me, you have to change classes and it's harder to keep track of things."*<br><br>*"I like tried [to get to bed earlier], but maybe some nights I would have homework or I'd be out late, so I couldn't really get on a schedule."* | *"... in the morning we get up, get – he gets a shower, has breakfast, and he gets ready and leaves for school, and there's not really anything that we could do differently to change that. You know, it's a set time that he's got to go, so there's not much we could change about the morning schedule."*<br><br>*"Um, but he often times will have them [electronics devices] on, which one of the – I have to admit that part of the struggle with the electronics is that school has, um, become clapping them to use electronics, because he's got homework every week that has to be done on a computer."* |

| | | |
|---|---|---|
| Competing extracurricular activities | *"I wouldn't really have much time to like do homework or eat a snack or anything because I have to go right to gymnastics. And um, and then at 8:45 so that's like when it ended. And then when I got home it was like usually like five minutes to 9:00 and I would have to come home, uh, still eat dinner and take a shower, and I would still have homework. So I wouldn't get to bed until like 10:30."* | *"Just the activities kind of happen when they happen so it's not really an option for her to sleep later."* |
| Long-term sustainability | *"It's hard because like now that the study is over I usually feel like I can just go back to what I am doing but its not good so I always try here and there so it just doesn't really work for me because I always feel like staying up"* | *Um, it was something we just did during the study. And since it ended I think we kind of – kind of gone back to our old habits. But I've noticed recently I've just been exhausted. Now that I'm thinking about it like I really should …try and get back on it. Um, just um, you know, especially with the uh, trying to lighten up on the – on the extracurricular."* |
| Suggested changes for future intervention research | | |
| Desire for intervention personalization | *"Um, well, I think that um, in the study there should have been more like the, um, things like uh, to have someone go to bed earlier, like when you were reporting your um, daily routine that they could, uh, figure out like maybe what was going on why you weren't getting enough sleep and then help you, not just like you know, tips buzzing on, um, your mom's phone, stuff like go to bed or watch T.V. one hour before sleeping."* | "*No I just think that, you know what maybe just in like if you find out what the child likes, like a favorite picture or something or little stickers, you can send little stickers I think you know that's helpful. Children are in to that so she likes the messages but maybe little pictures or something to kind of engage them even more I think that would help."* |

Note. * indicates theme reflected an intervention facilitator in some cases and an intervention barrier in others.
